# Harnessing Digital Phenotyping to Advance University Student Mental Health (Brightline): Study Protocol for a Prospective Observational Study

**DOI:** 10.1101/2025.07.31.25332472

**Authors:** Sakura Ito, Chin-Siang Ang, Onno P Kampman, Kartikeya Rokde, Thisum Buddhika, Creighton Heaukulani, Zhi-Wei Tan, Farrah Adystyaning Dewanti, Evelyn W.M. Au, Vivien S Huan, Robert JT Morris, Andy WH Khong, Andy Hau Yan Ho

## Abstract

**Introduction:** Mental health issues such as depression and anxiety are highly and disproportionally prevalent among university students. Beyond the academic rigor, stressors imposed by a new environment result in them being vulnerable to the onset and manifestation of mental health symptomatology. Leveraging smartphones and wearables for digital phenotyping capabilities is an innovative approach for monitoring and intervening in the mental health conditions of university students. This provides a unique opportunity to collect and identify digital and behavioral biomarkers, subsequently enabling the development of predictive models to identify university students at risk.

**Methods and analysis:** This study — Brightline— will employ an observational study design over a 6-month period, recruiting 500 students from a major public university in Singapore. Passive data collection will occur continuously throughout the monitoring period through a wearable device (Fitbit Charge 6) and smartphone sensors via the Brightline app, which utilizes a digital phenotyping data collection platform. Active data collection will consist of self-report questionnaires to be completed at the beginning of the study and follow-up assessments at one, three, and six months after. The passive and active data collected will be analyzed to identify the digital biomarkers associated with depression, anxiety, stress, loneliness, and affect among university students. Predictive models of these mental health issues will also be developed.

**Ethics and dissemination:** This study was approved by the Nanyang Technological University Institutional Review Board (IRB-2023-894). Findings from this study will be published in peer-reviewed journals and presented at academic conferences.

**Trial registration:** NCT06770075.

**Strengths and limitations of this study:**

- Brightline is a large-scale longitudinal study that aims to examine the mental health of university students over time using smartphones and wearables.
- This study has significant clinical implications, offering insights into the early warning signs and trajectories of university student’s mental health.
- These findings may inform the development of personalized and preventative interventions for integration into university mental health services.
- Challenges in adhering to active and passive data collection may lead to missing data and participant dropout, potentially affecting the development of predictive models.

## INTRODUCTION

Mental health issues are a significant and escalating public health concern, affecting approximately one in eight people worldwide [1]. Particularly troubling is the heightened vulnerability of young adults, who bear a disproportionate share of these challenges [2]. For many, university marks a critical transition into adulthood, yet this period is often fraught with health and behavioral concerns. The university experience is often accompanied by challenges across various domains, including separation from familiar social support networks [3], transitioning to a less structured environment, as well as pressures to achieve high levels of academic excellence [4]. Additionally, lifestyle changes, such as irregular sleep patterns and increased sedentary behavior [5], further exacerbate the mental health vulnerability of this demographic. Emerging evidence underscores the importance of this issue, with studies indicating that the onset of mental health disorders typically occurs before the age of 25 [6]. Research further reveals that the prevalence of depression and anxiety among university students can reach up to 85% and 55%, respectively—exceeding the rates observed in the general population [7, 8]. These mental health issues frequently coexist with other concerns, such as stress [9], loneliness [10], and negative affectivity [11].

Universities have generally established programs to promote student well-being, with a particular focus on mental health [12]. These initiatives traditionally emphasize treatment and intervention, including access to mental health services, well-being support, stigma reduction campaigns, and peer support programs [13]. The advent of digital interventions has further diversified the spectrum of available mental health support for university students [14, 15]. Despite these developments, help-seeking behaviors and the utilization of interventions among students remain low [16]. Barriers to seeking help include a lack of awareness about mental health symptoms, limited knowledge of available services, mental health stigma, doubts regarding the effectiveness of professional services, and a preference for self-reliance [17].

Beyond treatment and intervention, there is an urgent need to enhance the well-being of university students through health promotion [13]. A key aspect of this approach involves understanding the mental health profiles of both mentally healthy and mentally unwell students. Identifying these profiles can inform preventive mental health policies, thereby promoting the overall health of this population. In studies of university students’ mental health, typical assessment methods include clinician-administered or self-reported questionnaires. While these methods are validated for estimating the severity of mental health conditions, they have significant limitations, such as being time-intensive [18], inherently subjective [19], and unable to capture real-time changes in mental health states [20]. These assessments are often conducted intermittently, leading to significant gaps that may not reflect symptom changes over time. Moreover, they rely on retrospective self-assessment, making them susceptible to recall bias [21]. In addition, the scalability of these methods is limited, and it requires active involvement from individuals to complete [22]. An ideal assessment method would capture data continuously and minimize the effort required from students while remaining cost-effective.

Recent advances in digital phenotyping have revolutionized the monitoring and assessment of mental health states within individuals’ natural environments [23]. Smartphones and wearables can continuously and passively collect diverse behavioral, physiological, and environmental data beyond traditional self-reported surveys [24]. The integration of various sensors, such as accelerometers, gyroscopes, magnetometers, and GPS, has facilitated the ease of digital data collection. Data gathered in this fashion can be remotely gathered and used to identify digital and behavioral biomarkers— quantitative indicators derived from digital data that provide insights into an individual’s health status [25]. Examples of such biomarkers include heart rate, skin temperature, sleep patterns, and physical activity levels [26]. Using digital biomarkers derived from digital phenotyping to gain insights into the mental health state of university students has been a particular focus of recent studies. For instance, research has shown that digital biomarkers can predict depressive symptoms with 69.1% precision [27]. Additionally, during the COVID-19 pandemic, digital phenotyping was used to analyze passive smartphone data over a four-week period, revealing that sleep variance positively correlated with depression and stress scores [28]. Another study using smartphones demonstrated the ability to predict momentary fluctuations in anxiety symptoms using deep learning models [29].

Other studies have found significant correlations between smartphone usage and sensor data in predicting excessive stress levels among university students [30]. Furthermore, high evening screen time and overall phone usage have been associated with loneliness, with machine learning (ML) techniques achieving prediction accuracies of 82.43% [31], as well as the development of a loneliness model with an 82% accuracy rate by considering the impact of lifestyle and entertainment app features [32]. Such findings provide empirical support for the potential of digital biomarkers in screening and predicting mental health symptoms among students, guiding the prioritization and development of preventative measures and interventions [33–35].

Despite the promise of digital phenotyping, several limitations have hindered research progress. These include methodological limitations such as small sample sizes (Jacobson, 2019) or relatively short data collection periods [36]. A review study indicated an average of 81 participants and a 41-day study duration in digital phenotyping research [22]. There are also challenges with participant adherence, where ongoing efforts are needed to monitor participants and offer technical support to avoid issues of missing data that notably affect data quality [37]. Moreover, the issue of data quality, can significantly impact model accuracy and generalizability [38]. Such limitations introduce bias into datasets and subsequent development of algorithmic models [36, 39]. To address these challenges, prior studies recommend larger sample sizes, extended data collection periods, and robust data cleaning and preprocessing techniques [28]. However, conducting research in this area involves navigating a balance between the feasibility of methodological and data quality considerations.

Shorter studies with small samples may ensure better adherence but offer limited data. In contrast, longer studies with larger samples can offer more extensive and sufficient information for training machine learning models but face difficulties with missing data and participant adherence.

The current study utilizes the digital phenotyping app, Brightline, in a non-clinical setting of university students. The Brightline study uses the HOPES platform and is an application of its components toward assessing well and at-risk individuals, in the context of university student mental health. The HOPES platform, which was co-developed by the MOH Office for Healthcare Transformation and the Institute of Mental Health (a psychiatric care setting) in Singapore, was specifically designed to support digital phenotyping initiatives within both research and clinical settings [40]. Currently, the platform is being used to support clinical studies and services, particularly in the area of severe mental conditions [41–43]. It is built upon Beiwe, an open-source digital phenotyping platform [44, 45] and integrates data collection from wearable devices and smartphone sensors. Additionally, HOPES includes functionalities to streamline research processes, such as user-friendly onboarding, and dashboards to monitor participant adherence, ensure consistent data contribution, and maintain high data quality. The platform is designed with robust security measures to safeguard participant privacy and ensure secure data management and processing. It also offers data visualization and analytical tools to support exploratory statistical analyses and enable algorithm development. These features are also available in the Brightline app to enable the effective application of digital phenotyping toward developing predictive algorithms for university students.

### The present study

This study aims to address the limitations observed in past digital phenotyping research by focusing on the five V’s of big data: volume, variety, velocity, veracity, and value [46]. To tackle the challenges of small sample sizes and short data collection periods, we will collect an extensive volume of data from wearables and smartphones over a six-month period, involving a sample of 500 participants. To enhance data collection consistency and adherence, the Brightline app will centralize data collection from wrist-worn wearable devices and smartphone sensors. It will integrate a variety of data sources, including wearable and smartphone data, enabling real-time tracking and in-situ monitoring of participants’ daily behaviors to improve data velocity. To ensure data veracity, established protocols for data accuracy and quality control will be implemented, utilizing the data tracking dashboard to monitor data contributions. In addressing the value aspect of big data, this study applies machine learning (ML) algorithms for sophisticated and intricate analyses that are expected to outperform conventional methods in mental health prediction [47].

We aim to investigate the association between digital and behavioral markers, and mental health symptoms among university students through a multi-modal observational approach. This knowledge could help identify wellness indicators among university students, allowing university mental healthcare services and providers to better detect students who are at risk and connect them with relevant resources for effective intervention.

### Objectives

- To determine the digital and behavioral biomarkers associated with mental health issues—such as depression, anxiety, stress, loneliness, and affect —among university students.
- To develop predictive algorithms for mental health issues by identifying relevant digital biomarkers and training appropriate models using these features.
- To explore changes in mental health, university life event stressors, and help-seeking attitudes/behaviors among university students over time.
- To examine participant adherence to and acceptance of data collection protocols.

## METHODS AND ANALYSIS

### Study design

A prospective observational study will be conducted at a major public university in Singapore, monitoring participants over a six-month period. The study protocol has been registered on ClinicalTrials.gov (NCT06770075).

### Patient and public involvement

Patients and the public were not involved in the design, conduct or selection of outcomes in the study. Participant involvement was limited to the data collection of passive and active data collection across the study period.

### Participant recruitment and eligibility criteria

The study will recruit undergraduate students, a population at a critical developmental stage often characterized by significant academic, social, and emotional challenges [48, 49]. This demographic is well-suited for digital phenotyping research due to their high technology literacy and prevalent use of wearables and smartphones [50]. Multiple recruitment strategies will be employed, including university-wide email invitations, referrals through lecturers and recruitment briefings during a mandatory undergraduate course on healthy living and wellbeing. Interested participants will be directed to a Qualtrics form containing detailed study information and an embedded screening form to confirm eligibility.

To participate, individuals must meet the following criteria:

1. Be aged 18 years or older.
2. Be a current full-time undergraduate student.
3. Own a smartphone with Wi-Fi, 4G, and Bluetooth capabilities.
4. Possess adequate English language proficiency.
5. Be able to download the study apps.
6. Provide informed consent.

Individuals will be excluded if they:

1. Are part-time students.
2. Have a current diagnosis of any mental health disorder or a past diagnosis with any bipolar disorder, substance use disorder, or any psychotic disorder.
3. Are currently undergoing mental health treatment.
4. Lack sufficient English proficiency.
5. Report suicidal ideation as indicated by PHQ-9 Item 9, “Thoughts you would be better off dead or of hurting yourself in some way” [51].
6. Cannot commit to wearing a wearable device for the six-month monitoring period.
7. Receive special education accommodations or support from the university.

### Outcome measurements

Study outcomes will be assessed through both passive and active data collection methods [22], as illustrated in Fig 1. The schedule for outcome measurements is detailed in Table 1.

**Fig 1.**
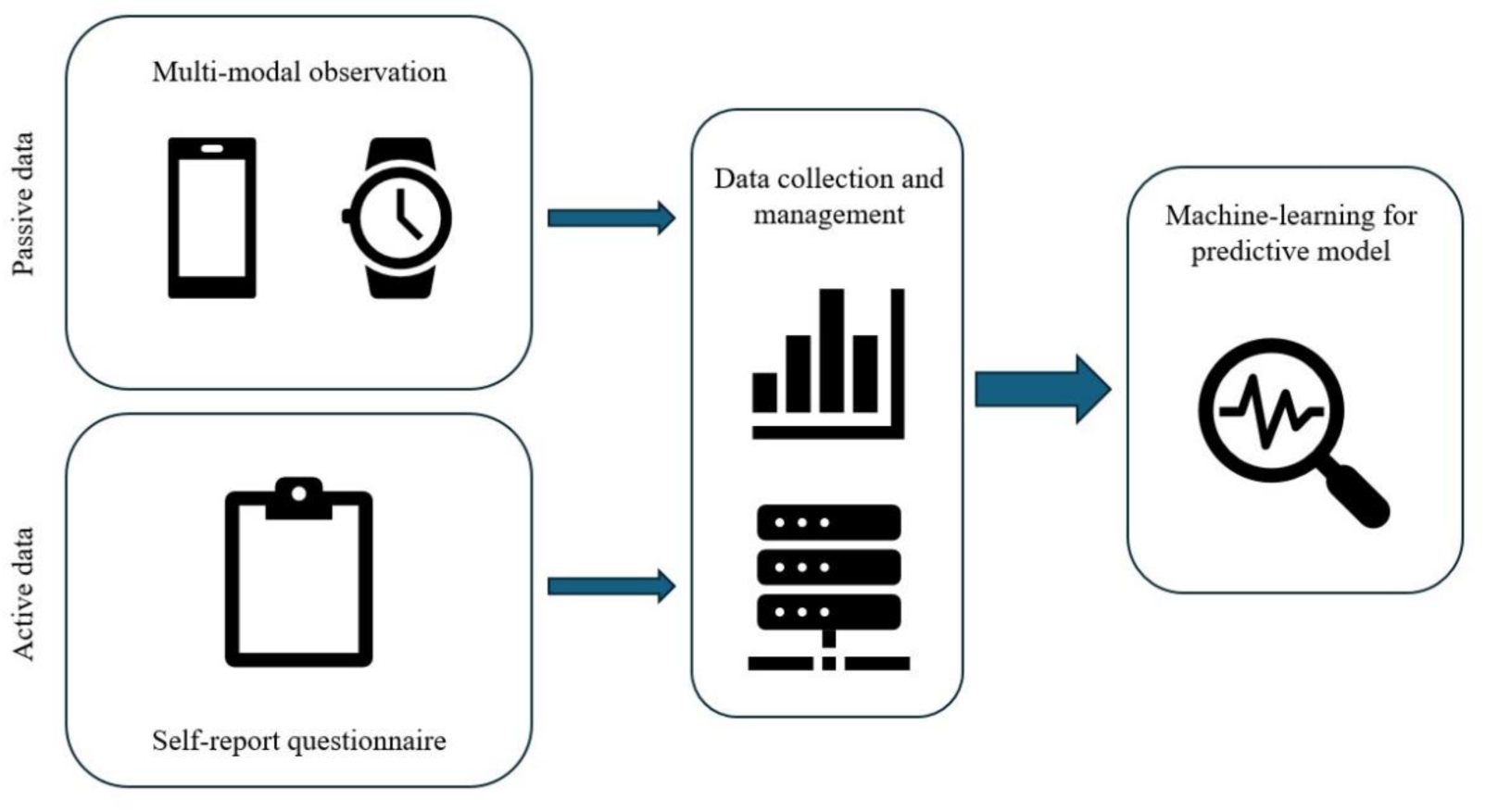
Multimodal data integration

**Table 1.**
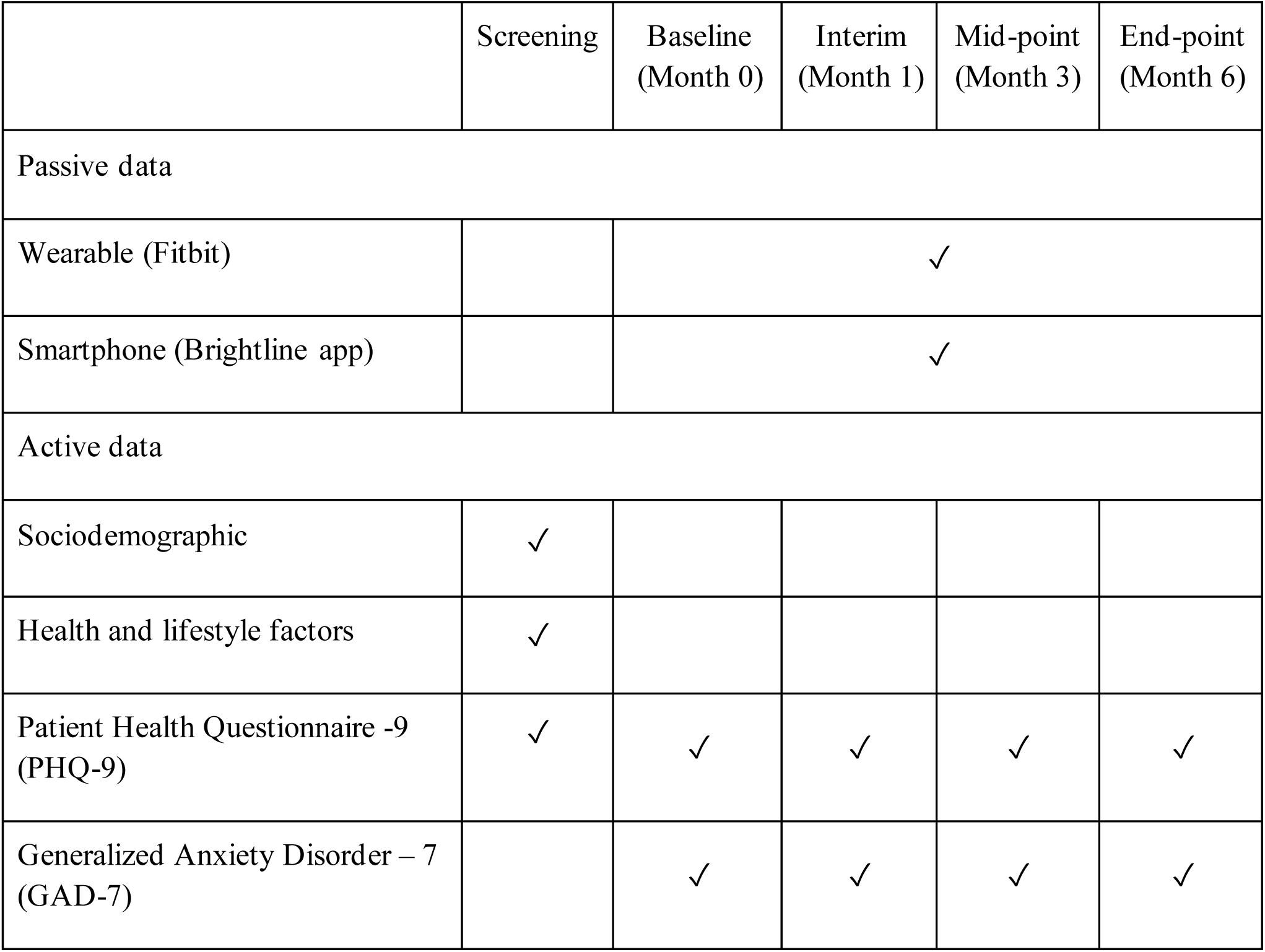
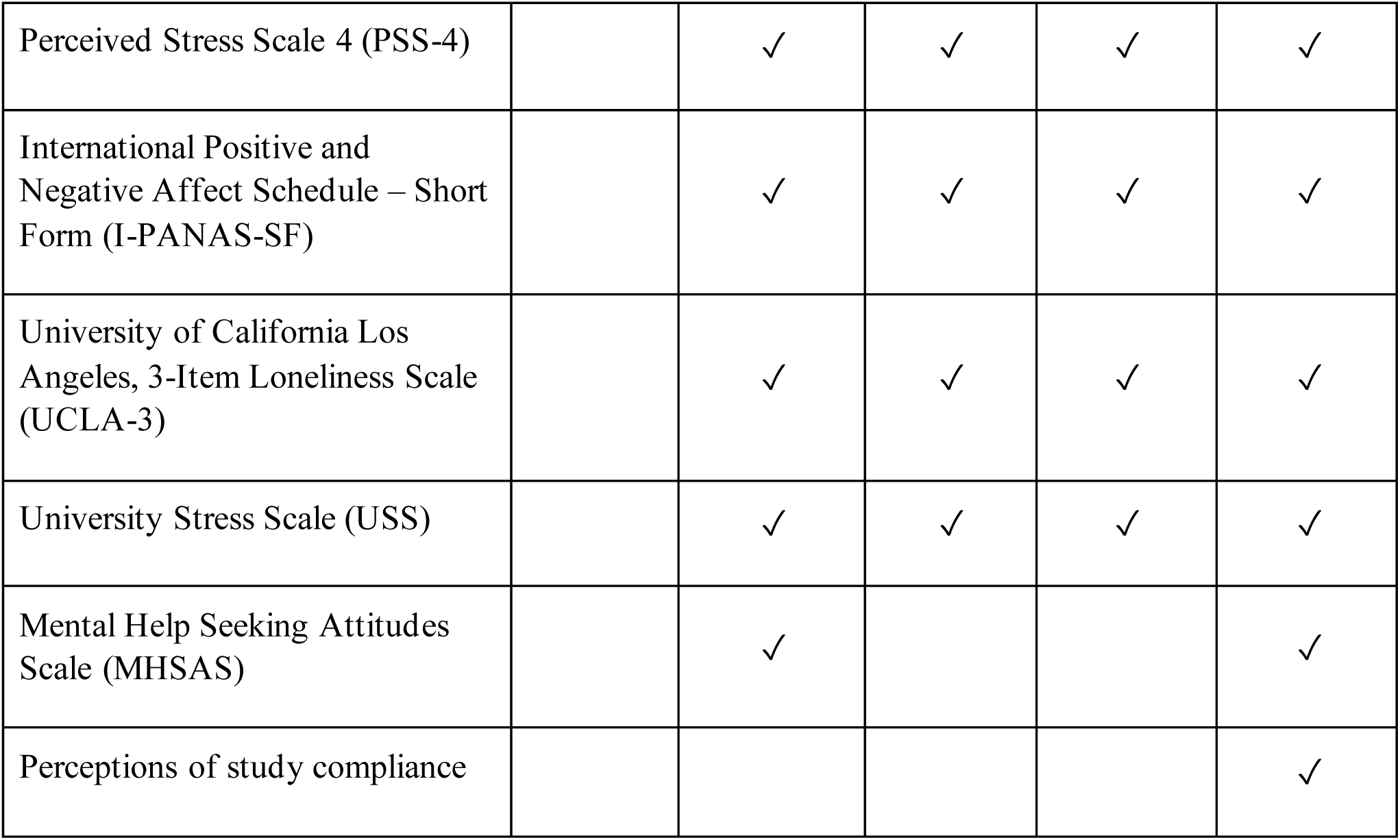
Data collection schedule.

### Passive data collection

Passive data will consist of digital phenotyping data collected via sensors embedded in a wrist-worn wearable device (Fitbit Charge 6) and a smartphone. This data will be continuously collected and transmitted to the Brightline platform; a dedicated app developed for this study (see Fig. 2 for interface).

**Fig 2.**
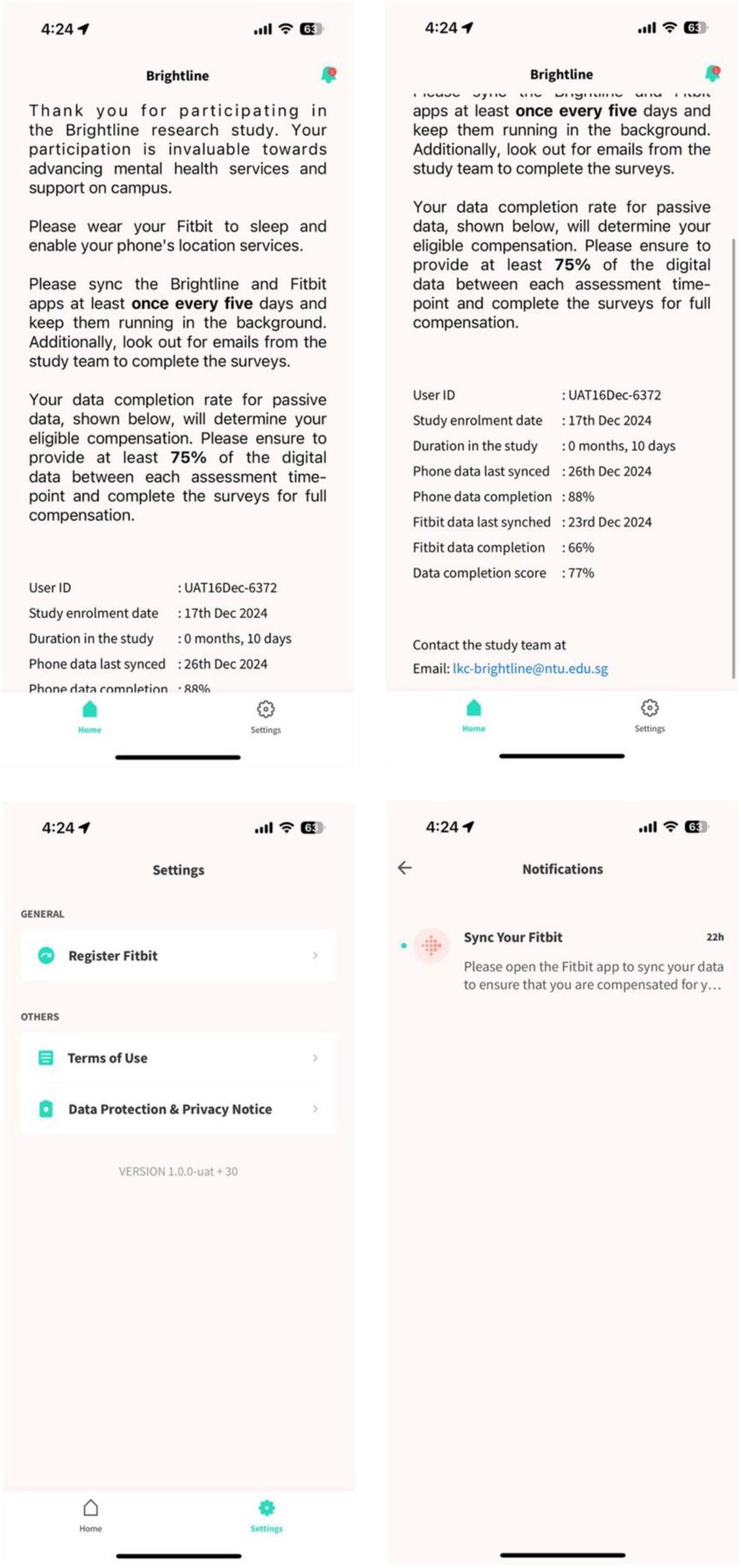
Brightline mobile app interface – Overview of the home screen featuring the data completion score, the settings page, and the notification page.

#### Wearable data (from Fitbit)

These categories of metrics are calculated using Fitbit’s proprietary algorithms.

1. Physical activity: Monitored using the embedded accelerometer sensor, which tracks activity patterns such as the number of steps.
2. Sleep: Detected by movement and heart rate metrics. Individuals are assumed to be asleep if no movement has been detected for an hour. Heart rate variability and movement is then analyzed to determine changes between sleep stages.
3. Heart rate and heart rate variability: Monitored using the embedded heart rate sensor, measuring beats per minute.

*Smartphone data (from Brightline app):*

The data collected may differ depending on whether participants are using iOS or Android for data collection.

1. Location: Tracked using GPS data to record locations, deviations from typical travel patterns, and variability in travel. Actual locations are obfuscated to ensure privacy.
2. Ambient light: Measured by a light sensor to detect the individual’s environmental context.
3. Phone states: Recorded by the app, which logs whether the screen is on or off, and the total time spent on the phone.
4. Finger taps: Logged by the app, capturing the number and timestamp of taps, screen orientation, and the apps interacted with. Specific keys typed are not tracked.
5. Sociability indices: Recorded by tracking the frequency and duration of calls and messages across platforms, as well as time spent on social apps. The content of communications is not recorded or accessed.

### Active data collection

Active data will be collected via self-reported questionnaires using Qualtrics (Qualtrics, Provo, UT), including:

1. Sociodemographic information. Participants will be asked the following information: age (in years), gender (male, female), ethnicity (Chinese, Malay, Indian, Others), family household monthly income level (Below $2000, $2000-3999, $4000-5999, $6000-9999, $10,000-14,999, $15,000 and above), student status (domestic student, international student), academic major, year of study (Year 1, Year 2, Year 3), cumulative academic performance (CGPA), and residence (off campus, on campus).
2. Health and lifestyle factors. Participants will be asked to provide the following information:

a. Health information: Body weight (in kilograms) and height (in centimeters). Self-rated health will be assessed by the question “In general, would you say that your health is:” with the responses categorized as “poor”, “fair”, “good”, “very good” and “excellent” [52].
b. Mental health: Participants will be asked of their family mental health history “Do you have any family history of mental-health related conditions?” (yes/no), and their help-seeking behaviors, “Have you used any mental health services in the past?” (yes/no).
c. Digital health: Participants will be asked: “Do you have any experience using digital mental health platforms/apps?” (yes/no), and “Do you have any experience with fitness trackers/wearable devices? (yes/no).
d. Lifestyle habits: Smoking behaviors (current smoker, non-smoker), alcohol consumption (yes/no) will be asked. Physical activity will be assessed by, “In a usual week, on how many days do you do vigorous activities (such as running, aerobics, etc.) for at least 10 minutes at a time, causing large increases in breathing or heart rate?” and “On days that you do vigorous activities for at least 10 minutes at a time, how much total time per day do you spend doing these activities?”[53]. Sleep duration will be based on, “On average, how many hours of sleep do you get in a 24-hour period?” [54], with responses categorized as “<6 hours,” “6 to 7 hours,” “7 to 8 hours,” “8 to 9 hours and “>9 hours.” Mobile screen time will be assessed by, “In the past month, how many hours on average per day do you spend looking at your mobile screen for entertainment or passing time?” with responses categorized as “<5 hours” or “≥5 hours [55].”
3. Mental health will be assessed exploring several dimensions, such as depression, anxiety, stress, positive and negative affect, loneliness.

a. Depression will be evaluated by means of the Patient Health Questionnaire-9 (PHQ-9). This is a nine-item scale measuring depression severity. It uses a four-point Likert scale from not at all to nearly every day. Total scores range from 0 to 27, with higher scores indicating increased depression severity [51].
b. Generalized Anxiety Disorder-7 (GAD-7) will be used to measure anxiety severity. This scale contains seven items, with a four-point Likert scale from not at all to nearly every day. Total scores range from 0 to 21, with higher scores indicating increased anxiety severity [56].
c. Perceived Stress Scale 4 (PSS-4). A four-item scale measuring stress experienced over the last month. It uses a 5-item Likert scale from never to very often. Total scores range from 0 to 16, with higher scores indicating increased stress levels [57].
d. International Positive and Negative Affect Schedule - Short Form (I-PANAS-SF). A ten-item scale divided into two subscales, measuring subjective experiences of positive and negative emotional states. It uses a five-item Likert scale from very slightly or not at all to extremely. Scores range from 5 to 25 on each subscale, with higher scores on both subscales indicating higher levels of positive or negative affect [58].
e. University of California, Los Angeles (UCLA) 3-Item Loneliness Scale (UCLA-3). A three-item scale measuring levels of loneliness. It uses 3-item Likert scale from hardly ever to often. Scores range from 3 to 9, with higher scores indicating higher levels of loneliness [59].
4. University Stress Scale (USS). A 21-item scale measuring the degree to which university life events are appraised as stressful, such as academic/coursework demands, friendships, or study/life balance over the past month. It uses a 4-point Likert scale from not at all to constantly. Scores range from 0 to 66, with higher scores indicating higher perceived stress [60].
5. Mental Help Seeking Attitudes Scale (MHSAS). A nine-item scale measuring attitudes toward seeking mental health help. It uses a seven-point scale to measure participant’s perceptions between negative and positive attitudes. Scores range from 1 to 7, with higher scores indicating more favorable attitudes [61, 62].
6. Perceptions of study compliance. Participants will complete a questionnaire to evaluate the acceptability of the study procedure at the end of the study. This will include five items rated on a 5-point Likert scale surrounding satisfaction with participation, degree of Fitbit interference in day-to-day life, degree of Fitbit comfort over the course over the study duration, degree of difficulty maintaining the Fitbit’s charge and using the Fitbit wristband, and degree of change in day-to-day activities caused by wearing the Fitbit [63]. Additionally, participants will be asked “Did you experience any additional difficulties or challenges throughout the study?”

### Study flow and data collection

Participants will follow the study flow presented in Fig 3. Passive data collection will occur continuously throughout the six-month study period, while active data collection will take place at four time points: Baseline (Month 0), Interim (Month 1), Mid-point (Month 3), Endpoint (Month 6).

**Fig 3.**
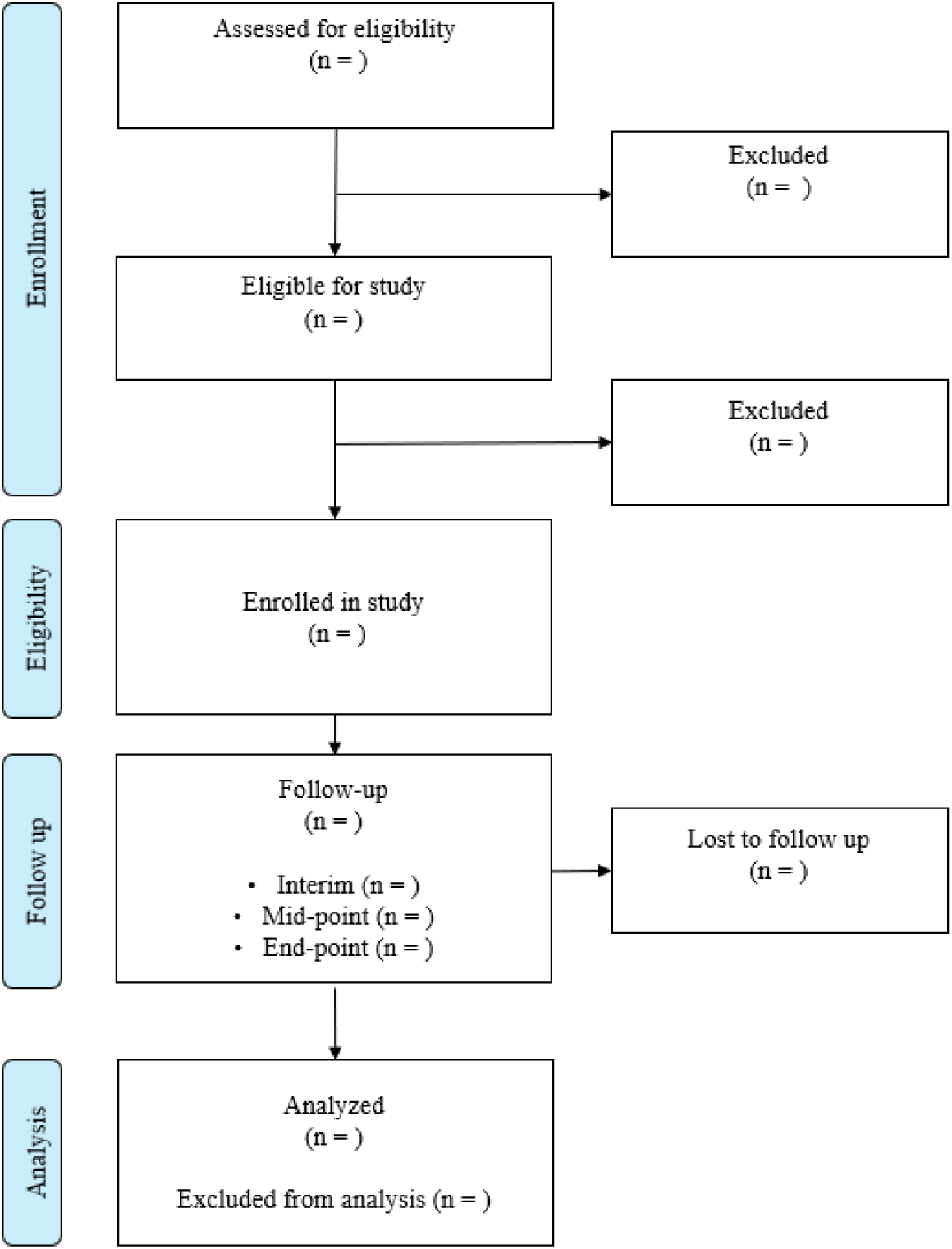
Study flowchart

#### Screening

Interested participants will first complete an online screening form to determine eligibility. Those who are deemed eligible and provide informed consent will proceed to the onboarding session.

#### Baseline (Month 0)

During the onboarding session, participants will receive the Fitbit device and be instructed to download the Fitbit and Brightline applications for data collection. The study team will assist participants in setting up the devices, logging into accounts, and synchronizing the app and devices. An overview of the study procedures and instructions on using the Fitbit and the Brightline app for data collection will be provided, with opportunities for participants to ask questions to the study team. At baseline, the first assessment of active data will be collected from the participants and passive data collection will be initiated. The active data will consist of self-reported questionnaires and will be administered remotely using Qualtrics.

#### Follow-ups (Month 1, Month 3, and Month 6)

Participants will complete three follow-up assessments at the interim (Month 1), mid-point (Month 3) and end-point (Month 6) of the study. Throughout the six-month study period, participants will be asked to wear the wearable device throughout the day except when bathing or charging the device.

They will be encouraged to access their Fitbit and Brightline dashboards to ensure successful data synchronization. Adherence to wearing the study device, granting necessary permissions, and maintaining active study app usage for continuous data synchronization will be self-monitored by the participants. Participants will be able to track their data contributions on the app home page. Nudges will be sent to participants through the app for additional reminders, with further support and reminders from the study team if necessary. For each follow-up, participants will complete the same questionnaire again. Upon the completion of six-month observation period, an off-boarding session will be conducted. During this session, the study team will debrief participants, retrieve the loaned Fitbit devices, and assist in uninstalling the study apps from participants’ mobile phones. Participants will receive compensation based on their engagement in both active and passive data collection methods. For active data collection, participants will be compensated for completing each survey.

Compensation for passive data collection will be based on the quantity of data contributed. Participants will receive the full allocated incentives if they provide at least 75% of the data captured by the wearable devices and smartphones between each assessment time-point, which follows the active data collection intervals.

### Ethical considerations and participant safety

Informed consent will be obtained remotely via the online survey platform Qualtrics prior to participation. Participants will be provided with a comprehensive informed consent form outlining the study procedures, compensation, rights, benefits, potential risks, and any discomforts associated with participation. The form will also include contact details of the study team to address any questions or concerns. A waiver of parental consent was obtained for participants aged 18 and older, as individuals under 21 are considered minors in Singapore. Participation in this study is entirely voluntary, and participants retain the right to withdraw at any point without incurring any penalties or adverse consequences. The confidentiality of all collected data will be rigorously maintained throughout the study. The collected data will only be accessible to members of the study team.

Given the sensitivity surrounding university student mental health, a referral network safety plan has been developed. Participants who exhibit moderate to severe symptoms of depression, anxiety, or stress—indicated by scores of PHQ-9 ≥10, GAD-7 ≥10, or PSS-4 ≥5 at any assessment point —will be referred to appropriate mental health resources. Students identified as at risk during the study will initially be provided with community helplines and support resources. To respect the autonomy of students in seeking mental health assistance, they will be asked if they are interested in accessing services through the university. For those who express willingness and provide consent, a formal referral will be made to the university’s counseling services for further assessment and intervention by trained professionals. This referral network, established in collaboration with the university’s counseling center, is designed to safeguard the well-being of students throughout the study.

Referred students will remain in the study and continue with data collection. However, if a participant indicates suicidal ideation on the final item of the PHQ-9 at any assessment point or exhibits displays any signs of suicidal ideation or self-harm to the study team, they will be immediately referred to university mental health services for emergency care. Following this referral, the participant will be withdrawn from the study and will undergo off - boarding procedures.

Additionally, if the study team assesses that a participant is experiencing circumstances that render them unfit or unable to continue, the participant will also be off boarded from the study.

### Sample size calculation

Given the absence of specific prevalence data on common mental health issues among students at our study site, we adopted a conservative approach to determine the sample size. We estimated a prevalence rate of 50% for our calculation [64, 65]. Using a 95% confidence level, a 50% expected prevalence rate, and a desired precision (margin of error) of 0.05, we calculated that a sample size of 384 participants would be required. To account for potential participant dropouts or incomplete data submissions, we incorporated a 30% non-response rate. This was informed by a study reporting a 30% discontinuation rate among users of fitness wearable technology within six months of acquisition [66]. This adjustment led to a target sample size of 500 participants, ensuring sufficient statistical power and accommodating potential data loss.

### Data analysis plan

#### Sociodemographic and baseline characteristics

Baseline characteristics of the participants and study variables will be reported using means and standard deviations or median and interquartile ranges for continuous variables, and frequencies in percentage for categorical variables. The associations between primary and secondary outcomes will be reported using Pearson’s or Spearman’s correlations.

#### Digital phenotyping analysis

Analyses will be conducted using the active data and the passive data. The raw passive data will be processed and cleaned, and any missing or invalid digital data will be removed. Digital biomarkers will be derived from the raw passive data collected from the wearable and smartphone. Exploratory analyses and review of previous literature associated with wearable and smartphone phone usage will be conducted to compute features that determine changes in mental health symptom severity and onset. We will assess associations between the self-report outcomes and digital phenotyping data features using correlation analyses using means after the study period and regression analyses. The digital phenotyping features may include those presented in Table 2 from passive data collection. ML algorithms will be trained and employed to predict mental health incidence and severity in undergraduate students. The statistical models that may be used include multi-level regressions, ensemble methods such as gradient boosting trees, deep learning methods including recurrent neural networks or transformers, and state-space models.

**Table 2.** Passive data features examples.

| Passive data collection method | Examples of data features |
| --- | --- |
| Wearable (via Fitbit) | 1. Physical activity – total steps, distance walked, calories burned, physical activity intensity, activity duration, inter-daily, sedentary activity, floors climbed, number of walks, duration of walks<br>2. Sleep patterns - time in bed, sleep efficiency score, sleep latency, wake after sleep onset, sleep stages, sleep offset timing<br>3. Heart rate and heart rate variability - heart rate beats per minute, heart rate variability, heart rate recovery, resting heart rate |
| Smartphone (via Brightline app) | 4. Location – time at home, number of locations, location entropy<br>5. Ambient light – intensity of light<br>6. Phone states – screen duration, phone sessions<br>7. Finger taps – number of taps, tapping speed, categories of apps tapped, unique apps used, typing errors<br>8. Sociability indices – duration and timing of calls, length and timing of messages sent and received, message count, time spent on certain apps |

Note. Due to the differences in the data collected, data features will depend on whether participants are using iOS or Android for data collection.

#### Data collection adherence and acceptability

The number and percentage of students completing the active data will be calculated for baseline, interim, mid-point and endpoint. For passive data, wearable adherence will be measured by calculating the proportion of days the wearable is worn over the six-month study period. The proportion of time within a day that the wearable is worn will also be evaluated. Similarly, smartphone adherence will be calculated by the proportion of days across the study period and the proportion of time within a day that smartphone data is available and collected. Additionally, participants’ response to the burden and acceptability of data collection procedures will be obtained to inform subsequent studies and/or interventional strategies.

### Study status

Subject recruitment commenced in January 2025 and data collection began in February 2025, with completion anticipated by September 2025. Preliminary results are expected by December 2025.

### Ethics and dissemination

Ethics approval for this study has been secured from the Nanyang Technological University Institutional Review Board (IRB-2023-894). The findings from this study will be disseminated through publications in peer-reviewed journals and presentations at academic conferences.

## SUMMARY

This study investigates the associations between digital behavioral and physiological markers and mental health symptoms among university students. Using a multi-modal observational approach, digital phenotyping offers a promising pathway to deepening our understanding of mental health within this population. The pervasive use of digital technologies in modern life allows data collected from wearables and smartphone sensors to provide a unique lens through which to explore the interaction between digital phenotyping and mental health. This research aims to develop digital biomarkers associated with mental health symptoms and construct predictive models that can be integrated into interventions within tertiary education settings.

Machine learning techniques will be applied to the collected data for pattern recognition and predictive analytics, aiming to enhance the early detection and monitoring of the onset and progression of mental health symptom. The derived models will elucidate both within-individual variability (e.g., temporal patterns in mental health behaviors) and between-individual variability (e.g., diverse manifestations of mental health behaviors). Such comprehensive analysis will provide a nuanced and precise understanding of mental health symptom changes at both the individual and group levels.

The significance of this study lies in its longitudinal, multi-modal digital phenotyping approach with a large sample size, offering a comprehensive view into the daily experience of university students and providing valuable insights into their mental health outcomes. However, the success of this study hinges on participant adherence to data collection, which may pose challenges. To mitigate this, we have carefully reviewed our data collection workflow and prepared a study implementation protocol aimed at enhancing adherence while minimizing missing data and dropout rates. This research will also contribute to the refinement of future digital phenotyping studies by assessing the feasibility and acceptability of conducting such research within this population and setting. Nevertheless, we acknowledge several limitations in our study sample. First, we exclude students with a current diagnosis of any mental health disorder or a history of bipolar disorder, substance use disorder, or any psychotic disorders. Therefore, this study focuses exclusively on the development and progression of mental health symptoms in students who are otherwise considered ‘well.’ Second, while the undergraduate student population comprises multiple nationalities, this study is conducted solely from a single institution in Singapore, and the findings may have limited generalizability.

Overall, this study aligns with ongoing initiatives to advance digital phenotyping for mental health and wellbeing, particularly in the measurement of mental health symptoms and personalizing mental health care [24, 47].

This observational study has the potential to significantly impact student mental health by offering valuable insights and a detailed characterization of mental health through longitudinal and comprehensive monitoring. The findings from this research could inform the development of more effective university mental health promotion and intervention strategies through the applications of digital phenotyping. In the long term, this study, along with its future iterations, is poised to revolutionize university mental health services by addressing critical components of prevention, detection, and management of student mental health.

## Data Availability

The data generated from this study will not be made publicly available due to its sensitivity.

## ACKNOWLEDGEMENTS

The authors would like to thank Malar Palaiyan for her support in the development of the referral network safety plan to ensure student safety during the study.

## AUTHOR CONTRIBUTIONS

SI and CSA drafted the manuscript with input from all co-authors. AHYH and AWHK secured funding and provided supervision. All authors were involved in one or more aspects of the study, including conceptualization, protocol development, or and have reviewed and approved the final version of the manuscript.

## DATA STATEMENT

The data generated from this study will not be made publicly available due to its sensitivity.

